# Evaluation of population immunity against SARS-CoV-2 variants, EG.5.1, FY.4, BA.2.86, JN.1, and JN.1.4, using samples from two health demographic surveillance systems in Kenya

**DOI:** 10.1101/2024.06.26.24309525

**Authors:** Doreen Lugano, Bernadette Kutima, Makobu Kimani, Antipa Sigilai, John Gitonga, Angela Karani, Donald Akech, Boniface Karia, Abdhalah K. Ziraba, Angela Maina, Arnold Lambisia, Donwilliams Omuoyo, Daisy Mugo, Ruth Lucinde, Joseph Newman, Dalan Bailey, Eunice Nduati, George Githinji, Charles N. Agoti, Philip Bejon, J Anthony G Scott, Ambrose Agweyu, Wangeci Kagucia, George M Warimwe, Charles Sande, Lynette I Ochola-Oyier, James Nyagwange

**Author notes:** Authors contributed equally (Doreen Lugano, Bernadette Kutima, Lynette I Ochola-Oyier, James Nyagwange). Corresponding author at KEMRI-Wellcome Trust Research Programme P.O. Box 230 Kilifi.

## Abstract

Increased immune evasion by emerging and highly mutated SARS-CoV-2 variants is a key challenge to the control of COVID-19. The majority of these mutations mainly target the spike protein, allowing the new variants to escape the immunity previously raised by vaccination and/or infection by earlier variants of SARS-CoV-2. In this study, we investigated the neutralizing capacity of antibodies against emerging variants of interest circulating between May 2023 and March 2024 using sera from representative samples of the Kenyan population. From our genomics data, we identified the most prevalent Kenyan and global variants and performed pseudoviruses neutralization assays with the most recent SARS-CoV-2 variants. Our data show that antibodies from individuals in the general population in Kenya were less effective against the recent prevalent SARS-CoV-2 omicron variants (i.e. EG.5.1, FY.4, BA.2.86, JN.1, and JN.1.4) compared to the ancestral wildtype strain. Although there was increased neutralization following multiple doses of vaccine, antibodies from >40% of the vaccinated individuals did not neutralize the omicron variants, suggesting that individuals were susceptible to infection by these variants.

## Introduction

The emergence of new variants, recurring natural infection, vaccine efficacy, and specificity and longevity of neutralizing antibodies are key variables determining long-term control of coronavirus disease 2019 (COVID-19) ^1,2^. Research efforts have focused on finding interventions that can reduce infection and transmission while improving clinical outcomes of COVID-19 in patients ^3^. A main strategy is the raising of protective neutralizing antibodies (nAbs) in the host population through vaccination ^4–7^. Neutralizing antibodies bind to SARS-CoV-2 structural proteins, especially the spike, inhibiting infection of host cells through the ACE-2 receptor ^8–10^. However, the virus continues to mutate in the spike protein giving rise to new variants. This makes it unclear to what extent neutralizing antibodies induced by earlier exposure through natural infection and vaccination are protective to the emerging variants and whether mono-variant multiple vaccine dosing improves this outcome.

The first vaccinations in Kenya began in adults in March 2021, and as of May 2022, the Ministry of Health reported approximately 8.3 million fully vaccinated adults and 2.5 million partially vaccinated adults ^11^. This accounted for only 30.7% of the Kenyan adult population being vaccinated and highlights issues with access, vaccine hesitancy due to religious and cultural beliefs, and concerns over safety and efficacy ^11,12^. Several studies report on vaccine effectiveness in the rapid production of neutralizing antibodies against SARS-CoV-2 ^1,2,8–10,13^ and similarly, natural infection leads to the development of wildtype and cross-protective nAbs within the first two weeks after infection ^1,2^.

Emerging SARS-CoV-2 variants have increased mutations targeted to the spike gene, that lead to immune escape phenotypes ^4,6^. Questions have been raised on whether ancestral strain COVID-19 vaccines are sufficient to counter upcoming variants of concern and interest, and new strategies on access and composition of vaccines are fast changing ^4,14^. Alternatives such as boosting with monovalent vaccines comprising more recent variants may improve neutralizing antibody function and outcomes of COVID-19 in patients and the World Health Organization Scientific Advisory Group of Experts (WHO-SAGE) recently recommended the development and deployment of JN.1 spike-based vaccines ^14–17^.

Here we used genomic data to identify locally circulating variants in Kenya between May 2023 and March 2024 ^18,19^. We evaluated the neutralizing capacity of antibodies from samples collected from two health demographic surveillance systems in Kenya (n=58), for vaccine-induced immunity against dominant variants EG.5.1, FY.4 ^19^, BA.2.86, JN.1, and JN.1.4. As a comparator, we used natural wildtype infection samples (n=20) collected in 2020 at the onset of the pandemic and confirmed to be PCR positive for SARS-CoV-2. We also assessed the effects of different doses of vaccine, age, sex, and type of vaccine on antibody neutralizing titers.

## Results

### Circulating SARS-CoV-2 lineages between May 2023 and March 2024

We used genomic data to identify emerging variants of interest in Kenya and globally, between May 2023 and March 2024 ^21^. The top circulating strains worldwide were XBB.1.5, JN.1, JN.1.4, BA.2.86, EG.5.1.1, (Fig. 1A) whereas XBB.1.5, XBB.1.16, JN.1, JN.1.4, and BA.2.86, were prominent in Europe (Fig. 1B). Kenya’s leading strains were FY.4 ^19^, GE.1.2, JN.1, JN.1.4, and JE.1.1 (Fig. 1C). We mapped 616 sequences from samples collected and sequenced in Kenya in a phylogenetic tree. These samples were collected in 17 counties, with the majority being from Nairobi (28%), Kilifi (20%), and Kiambu (13%) counties (Supplementary Fig. 1). We show the predominance of strains FY.4 ^19^, GE.1.2, JN.1, JN.1.4, BQ.1.1, and JE.1.1/KH.1 like variants (Fig. 1D). We performed pseudovirus neutralization assays on circulating strains EG.5.1, FY.4, BA.2.86, JN.1, and JN.1.4. FY.4, JN.1, and JN.1.4 were prevalent in Kenya during this period ^18,19^, while BA.2.86 and EG.5.1 were circulating in nearby regions ^25–27^.

**Figure 1:**
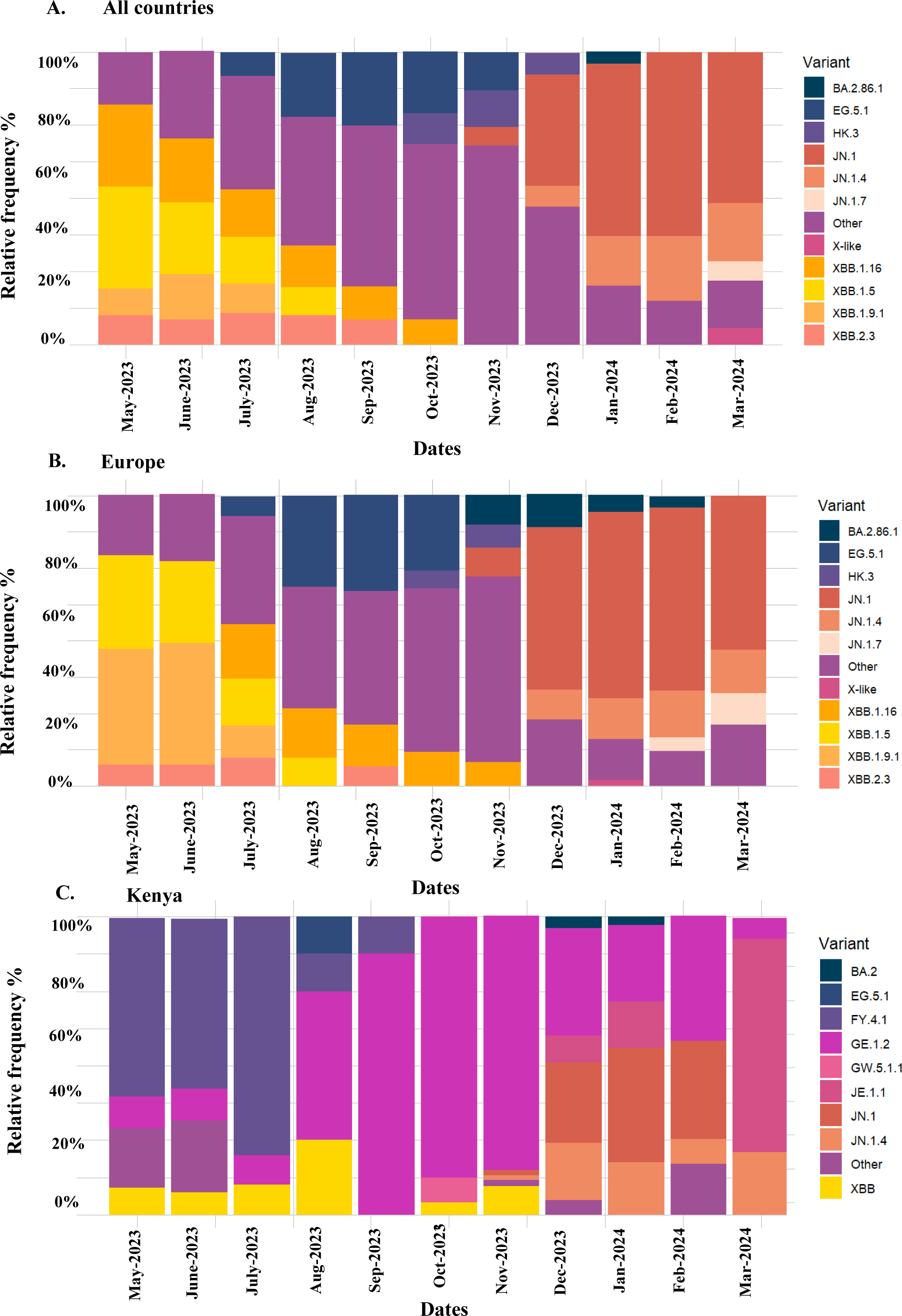
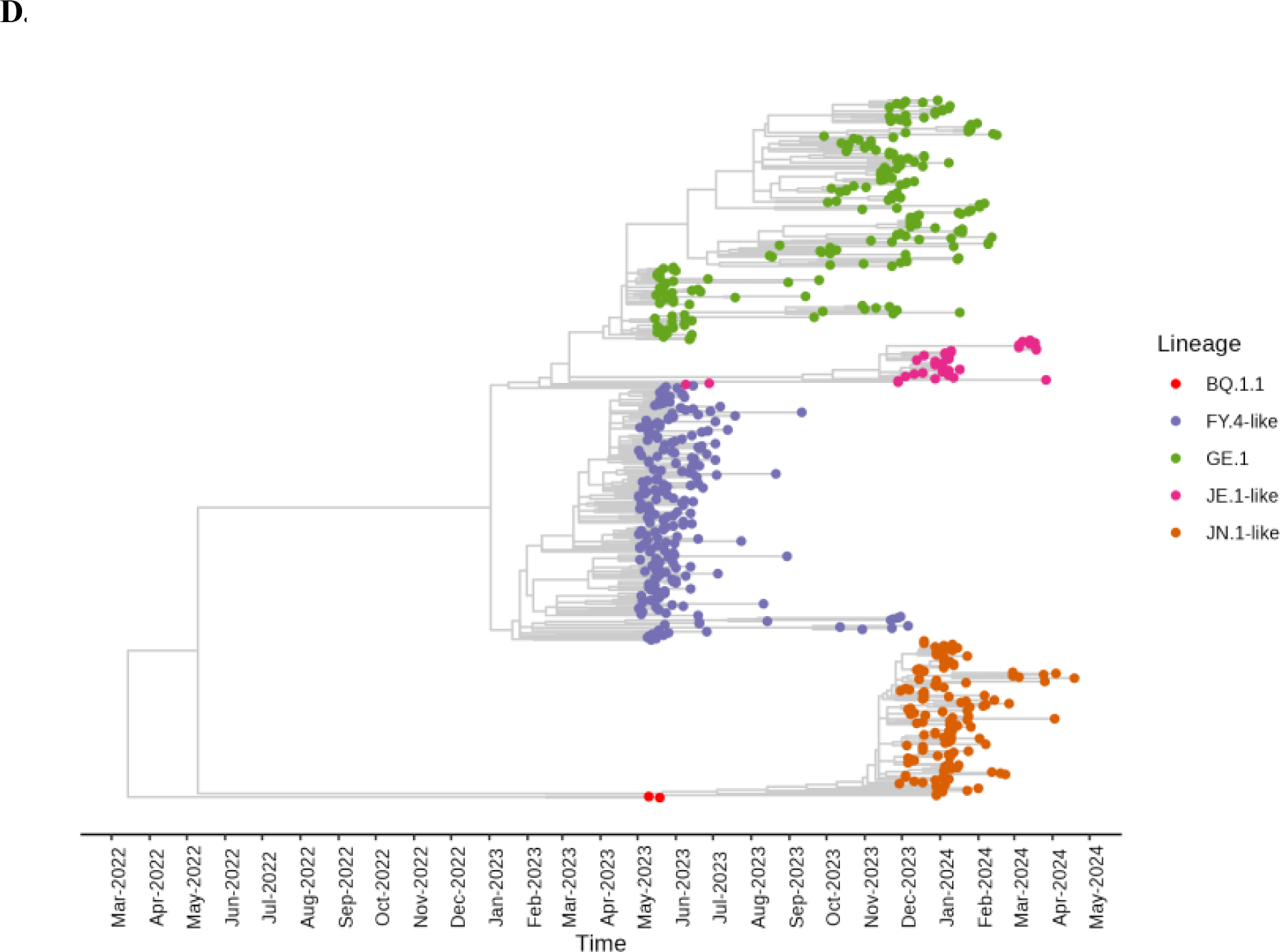
SARS-CoV-2 lineage frequency (%) between May 2023 and March 2024. This figure presents worldwide, European, and Kenyan SARS-CoV-2 lineage frequency % maps from GISAID. A. Shows the lineage frequency % in all countries (n =775112) B. Shows the lineage frequency % in Europe (n=232081). C. Shows the lineage frequency % in Kenya (n=616). The legends in A, B, and C show the top circulating variants in each region within the period of interest, with the colors corresponding to the frequency. D. Shows the phylogenetic analysis of 616 sequences collected and sequenced from Kenya between May 2023 and March 2024. The legend shows the lineages corresponding to the colors on the tree.

### An evaluation of neutralizing function against, wildtype, EG.5.1, FY.4, BA.2.86, JN.1, and JN.1.4, in population-representative samples

Inhibitory dilutions (ID_50_) of sera from naturally infected (n=20) and vaccinated individuals (n=58) were used to determine neutralizing antibody function against EG.5.1, FY.4, BA.2.86, JN.1, and JN.1.4 pseudovirus variants. Compared to wildtype pseudovirus where (83%) 65/78 serum samples exhibited neutralization function, only 41/78 (53%) neutralized EG.5.1, 23/ 78 (29%) neutralized FY.4, 25/78 (32%) neutralized BA.2.86, 29/78 (37%) neutralized JN.1, and 31/78 (40%) neutralized JN.1.4 (Fig.2A-E). Furthermore, among the positives we observed a consistent decline in neutralizing antibody titers against all strains with a mean ID_50_ of 1:613, 1:441, 1:576, 1:530, and 1:532 for EG.5.1, FY.4, BA.2.86, JN.1, and JN.1.4, respectively, compared to 1:1155 in wildtype (Fig.2A-E). These differences in ID_50_ of the variants relative to wildtype were statistically significant (p-value <0.0001, Fig.2F).

**Figure 2:**
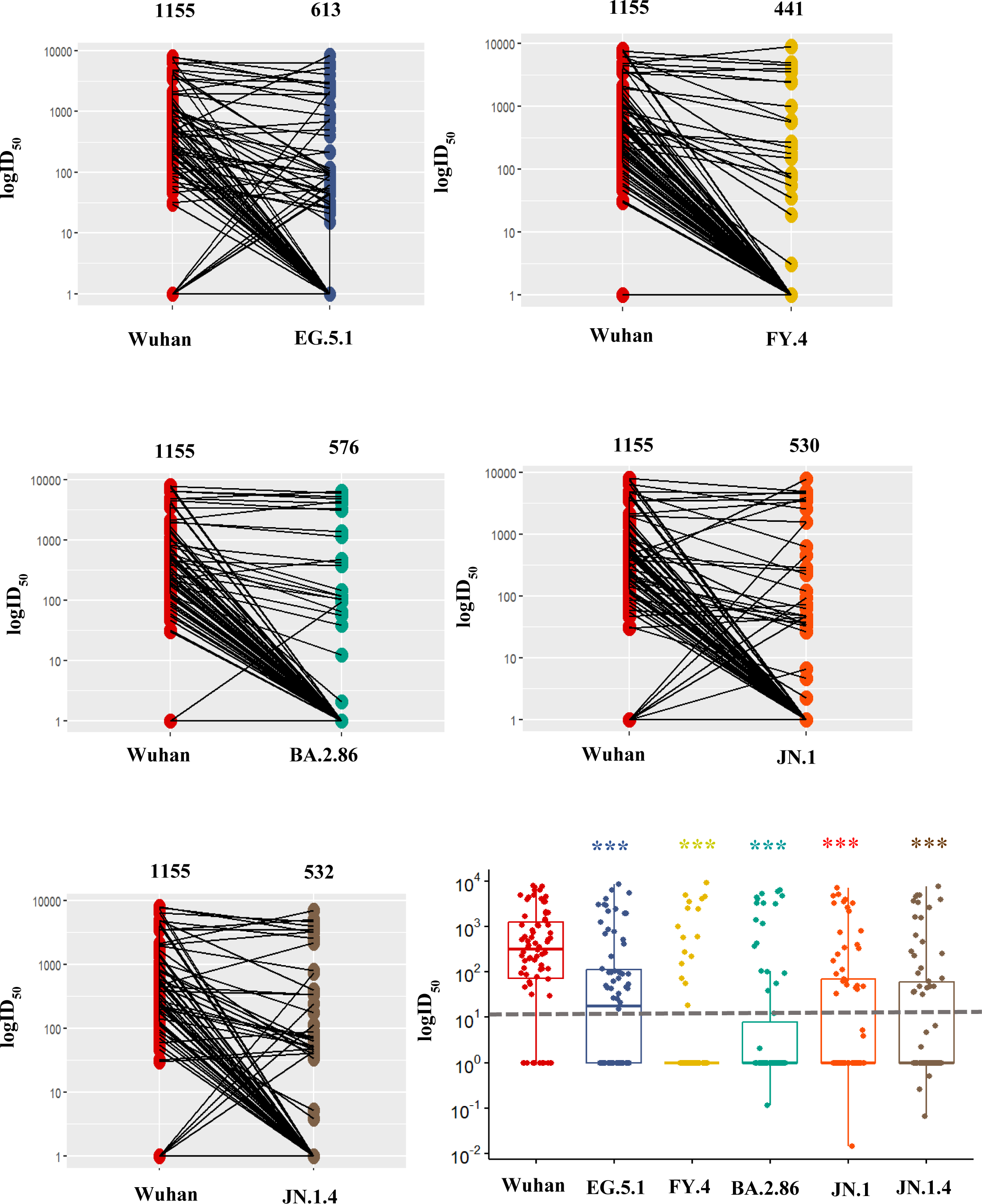
A comparison of inhibitory dilutions (ID_50_) between wildtype and omicron variants EG.5.1, FY.4, BA.2.86, JN.1, and JN.1.4, in Kenyan population samples (n=78) A. Represents the log ID_50_ between wildtype and EG.5.1 where 41/78 in EG.5.1 showed neutralization. B. Represents the log ID_50_ between wildtype and FY.4 where 23/78 samples showed neutralization. C. Represents the log ID_50_ between wildtype and BA.2.86 where 25/78 samples showed neutralization. D. Represents the log ID_50_ between wildtype and JN.1 where 29/78 samples showed neutralization. E. Represents the log ID_50_ between wildtype and JN.1.4 where 31/78 samples showed neutralization. The numbers above each variant plot represent the mean ID_50_ compared to wildtype. F. Shows the statistical analysis of ID_50_ between wildtype and each variant. A statistical significance of P< 0.0001 (***) was recorded between wildtype and all strains, based on Mann-Whitney tests. The sample colors represent SARS-CoV-2 variants where red is wildtype, blue is EG.5.1, yellow is FY.4, green is BA.2.86, orange is JN.1, and brown is JN.1.4. The dotted line represents the cut-off neutralization ID_50_ of 10^1^.

Next, we measured the escape from the existing humoral immunity by these variants after natural infection with the wildtype variant and vaccination with different doses. Neutralization in samples with wildtype natural infection showed that 18/20 (90%) individuals could neutralize the wildtype but only 4/20 (20%) neutralized EG.5.1, 3/20 (15%) neutralized FY.4, 3/20 (15%) neutralized BA.2.86, 3/20 (15%) neutralized JN.1, and 3/20 (15%) neutralized JN.1.4 (Fig.3A). After one dose, there was increased neutralization of 12/20 (60%) in EG.5.1, 5/20 (25%) in FY.4, 4/20 (20%) in BA.2.86, 8/20 (40%) in JN.1, 7/20 (35%) in JN.1.4, and 15/20 in wildtype (75%). However, after three doses, there was significant decrease in neutralization function against most omicron variants relative to two doses, where 12/19 (63%) individuals for EG.5.1, 3/19 (16%) for FY.4, 7/19 (37%) for BA.2.86, 7/19 (37%) for JN.1, and 8/19(42%) for JN.1.4 (Fig.3B).

**Figure 3:**
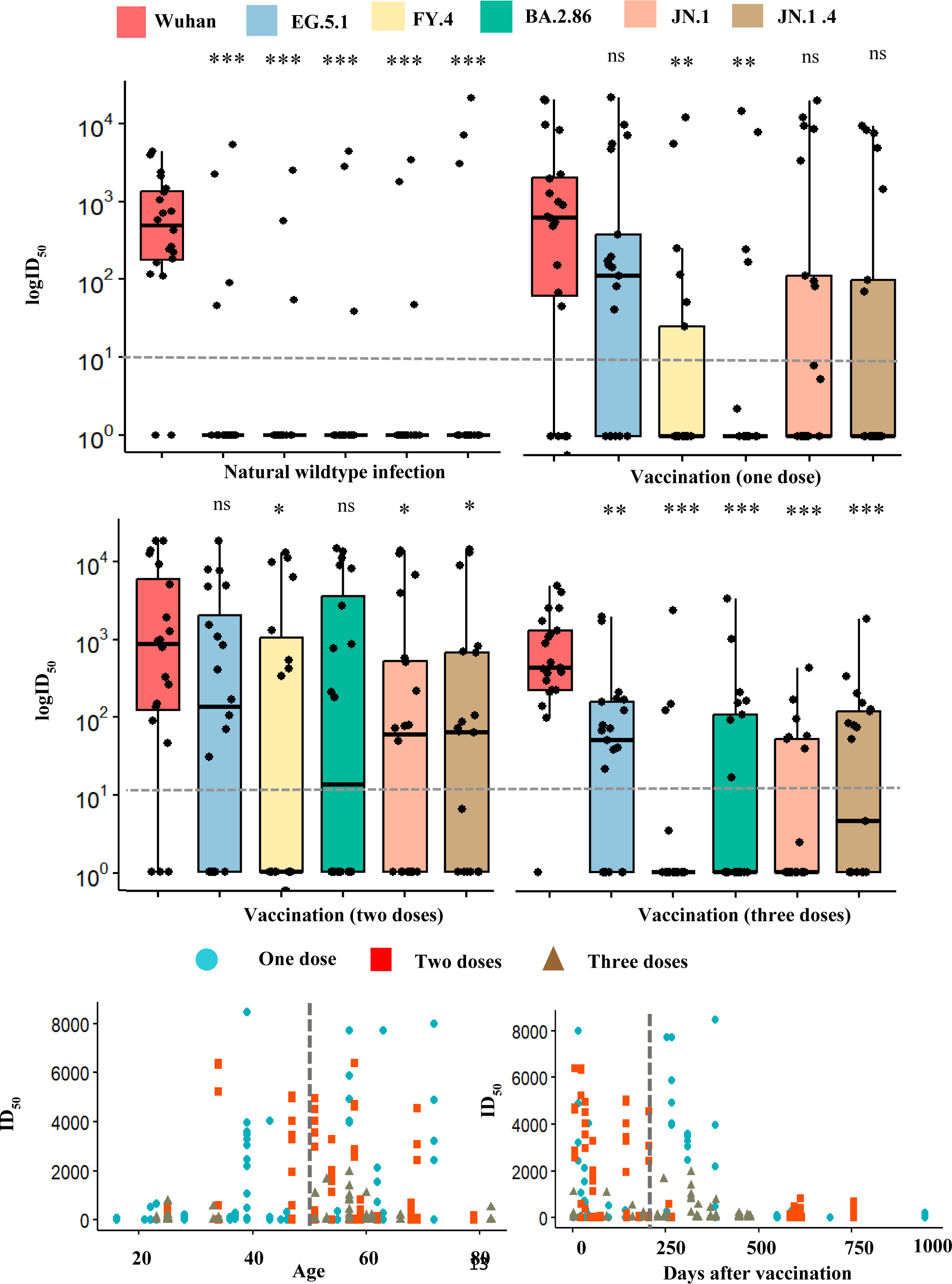
Evaluation of naturally-acquired (n=20) and vaccine-induced immunity (n=58) in Kenyan population samples. A & B. Shows neutralization (log ID_50_) after natural infection (n=20) or vaccination with one dose (n=20), two doses (n=19), and three doses (n=19). The legend represents SARS-CoV-2 variants where red is Wuhan, blue is EG.5.1, yellow is FY.4, green is BA.2.86, orange is JN.1, and brown is JN.1.4. Statistical significance was determined by Mann-Whitney tests where P<0.05 (*), P<0.001 (**), and P<0.0001 (***). The dotted line represents the cut-off neutralization ID_50_ of 10^1^. C. Represents the distribution of ID_50_ based on age between individuals administered with one, two, and three doses of vaccine. The dotted line separates individuals below and above 50 years old. D. Shows the distribution based on days post-vaccination with administering one, two, and three vaccine doses. The dotted line represents a cut-off of 6 months after vaccination. C & D Circles represent one dose, boxes represent two doses, and triangles represent three boxes.

We then examined whether the age of individuals who received one, two, or three doses of the vaccine affected these findings. From the distribution of ages, we noted that the majority of individuals who received two doses 14/20 (70%) and three doses 14/19 (74%) were above 50 years (Fig. 3C). Additionally, we determined whether days post-vaccination affected variances in neutralization function. We found that 9/20 (45%) of individuals receiving two doses were within 6 months of vaccination (180 days), whereas only 3/19 (15%) of individuals with three doses were within this period, which could explain the decline in neutralization (Fig.3D). Lastly, we evaluated the neutralization capacity between male and female participants and the type of vaccine administered and observed no markable changes (Supplementary Fig. 2)

## Discussion

The rapid mutation of the SARS-CoV-2 virus may lead to the emergence of new variants that evade neutralization by pre-existing antibodies and have increased infectivity, transmissibility, and pathogenicity ^28–32^. This study investigated neutralization as a correlate of protection against emerging variants circulating between May 2023 and March 2024 in a subset of the Kenya population. We used genomic data from Kenya ^18–20^, to identify variants circulating during the study period and performed pseudoviruses neutralization assays.

Genomic data from samples collected in 17 counties in Kenya (Supplementary Fig. 1) showed mostly similar lineage frequencies compared to other regions globally, but there were occasional differences such as predominant circulation of FY.4 and GE.1.2 in the Kenyan population only (Fig. 1 A-C) ^19,21^. We characterized EG.5.1, FY.4, BA.2.86, JN.1, and JN.1.4. FY.4 was first reported in March of 2023 and was prevalent in Kenya, specifically in the coastal region, with a Y451H mutation in the spike and P42L in the ORF3a (Fig. 1C-D) ^19^. Functional changes caused by Y451H remain unknown, but mutations on the ORF3a are linked to loss of CD8+ T cell recognition ^19^. EG.5.1 is a descendant of XBB.1.9.2 lineage which emerged in May 2023 and demonstrated substantial growth advantage over predecessor XBB lineages in Europe, Asia, and North America ^25^. BA.2.86 was a variant of interest with more than 30 mutations on the spike protein relative to ancestral strain BA.2 and had over 35 mutations compared to XBB.1.5 ^26,33^. It was first reported in mid-July 2023 and was highly prevalent in Israel, Europe, and the US, and as of November 2023, it was detected in Kenya (Fig. 1A-D). Lastly, JN.1 and JN.1.4 are sub-lineages of BA.2.86 and maintain the hallmark L455S amino acid change on the spike and three key mutations in non-spike regions ^25^. These two later strains were the most recent topmost circulating variants in many global regions and were reported to enhance immune evasion properties and increase infectivity ^25^.

Surveillance of neutralizing capacity in a subset of population samples allows insights into the general immunity and vaccine effectiveness, enabling the generation of better strategies to mitigate future outbreaks ^15,34^. We observed that most wildtype-induced infections elicited antibodies that could not neutralize the new variants. Interestingly, a few of these antibodies showed neutralization capabilities against the emerging omicron variants, suggesting the presence of cross-protective antibodies and presenting an opportunity for the isolation of cross-protective monoclonal antibodies for therapeutic use, or inform the design of vaccines that specifically induce cross-reactive antibodies.

We have demonstrated that both natural infection and vaccine-induced immunity from the wildtype was neutralizing to the wildtype but non-neutralizing against the emerging omicron variants. Although there was a substantial boost to neutralizing antibody titers against all new variants after two doses, there was no evidence of further boosting after three doses, and possibly even reductions against some variants. Similar observations have been reported in sera from 3-dose mono-valent vaccinated individuals who exhibited non-neutralizing antibodies against BA.2.86, and XBB variants ^26^. However, in our study cohort, 12/19 (63%) individuals with three doses were above 50 years, and 16/19 (84%) were sampled about 9 months (287 days) post-vaccination. Previously, it has been shown that for older individuals the level of protection waned by more than 50 % after three or more doses within 6-9 months ^33^. Therefore, the reduced neutralization after three doses observed in this study could have been confounded by the age of the cohort, and the longer sampling frame post-vaccination, > 9 months. Nevertheless, the data does not support repeated boosting as a strategy to generate cross-reactive antibodies.

Genomic surveillance of SARS-CoV-2 has greatly reduced in Kenya, hence all genomic data in this study represents two-thirds of all geo-specified isolates came from two counties of Kilifi and Nairobi, presenting a limitation of the study in generalization of the Kenya genomic data. Another limitation of this study is the use of convenient cross-sectional samples which implies the lack of follow up with participants to understand how pre and post-vaccination infections could affect the neutralization capacity. Therefore, the data presumed as vaccine-induced immunity could be caused by hybrid immunity and the differences could be driven by infections by different variants. Some studies have suggested that vaccination and boosting by the wildtype may result in immune imprinting and requirement for multiple boosting with omicron based vaccines to achieve neutralization of the new variants ^16,35^. However, it is not clear if natural infection could break the imprint to provide protective hybrid immunity. Also unclear is the impact of such imprinting to countries such as Kenya which have majorly administered mono-valent wildtype based vaccines and boosters. Nonetheless, mono-valent studies using omicron and its emerging subvariant boosters are in progress and may provide clarity in the future ^14^.

In conclusion, we demonstrate a decline in naturally acquired and vaccine-induced antibody protection, with the emergence of new omicron variants in a subset of the Kenyan population. This conclusion prompts the need for updated vaccine strategies in the country such as boosting by vaccines with currently circulating variants, to counter immune escape as the virus evolves, giving the population a chance to raise protective neutralizing antibodies to circulating variants ^14,15,16,34^. Although we have measured nAbs as a correlate of protection, other arms of the immune system may also play a role in the protection against COVID-19, such as T-cells ^36,37^. Furthermore, the absence of nAbs may not necessarily mean absence of memory B-cells which could be quickly mobilized to produce nAbs in case of an infection thereby conferring protection ^38^.

## Materials and Methods

### Genomics data

Genomics surveillance data from the Kenya Medical Research Institute Wellcome Trust Research Programme (KWTRP), and other sequencing facilities in Kenya were used to determine locally circulating variants between May 2023 and March 2024 ^18–20^. The data included samples collected from 17 Kenyan counties and sequenced in four Kenyan facilities, KWTRP (57%), Kenya Medical Research Institute-Center for Disease Control and Prevention (KEMRI-CDC) (39%), International Livestock Research Institute (ILRI) (2%), and National Public Health Laboratory (NPHL) (1%). The majority of sequences were from the Nairobi (28%) and Kilifi Health and Demographic Surveillance System (20%) ^18–20^ and Kiambu (13%) (Supplementary Fig. 1). Percentage of SARS-CoV-2 lineage frequencies for circulating variants in Kenya, Europe, and worldwide collected during the period of interest was based on data from the Global Initiative on Sharing Avian Influenza Data (GISAID) as of 4^th^ June 2024 ^21^. The sequences collected and sequenced in Kenya during this period were downloaded in a multifasta file and used to construct phylogenetic trees ^18,19^.

### Sample sets

To evaluate population immunity against the emerging variants, we took advantage of residual samples collected in the Kenya Multi-site Integrated Sero-surveillance study in the period of September to December 2022 (n=30) and July to October 2023 (n=30) ^22,23^. This study was approved by the Scientific Ethics Review Unit (SERU) under identification numbers 4085 and 4807. An inclusion criteria was the receipt of vaccination. We selected samples with well-documented information on vaccination such as a vaccination certificate or a short message service received after vaccination. As a comparator, we assayed plasma from 20 vaccine naïve SARS-CoV-2 wildtype naturally exposed individuals with PCR-positive results and sampled ≥7 days after their PCR-positive diagnosis ^17^. The vaccinated post-pandemic panel consisted of 58 individuals sampled from the Kilifi and Nairobi Health and Demographic Surveillance System (HDSS) site ^22^. The HDSS participants who were confirmed for vaccination were further split according to the number of doses received. A final classification of either one (n=20), two (n=19), or three doses (n=19) was defined and used for assaying. Detailed demographic characteristics of participants are shown in Table 1.

**Table 1:** Baseline characteristics of naturally infected and vaccinated cohort used in this study. The table provides details of the sex, age, vaccination status, and type of vaccine in this cohort.

|  | AstraZeneca<br>(n=30) | Pfizer<br>(n=10) | Johnson &<br>Johnson<br>(n=10) | Moderna<br>(n=6) | Mixed<br>(n=2) | Wildtype<br>Infected<br>(n= 20) | Total<br>(n=78) |
| --- | --- | --- | --- | --- | --- | --- | --- |
| **Sex** |  |  |  |  |  |  |  |
| Male | 15 (50%) | 2 (20%) | 5 (50%) | 3 (50%) | 0 (0%) | 11 (55%) | 36 |
| Female | 15 (50%) | 8 (80%) | 5 (50%) | 3 (50%) | 2 (100%) | 9 (45%) | 42 |
| **Age** |  |  |  |  |  |  |  |
| >18-60 | 24 (80%) | 8 (80%) | 8 (80%) | 5 (83%) | 2 (100%) | 17 (85%) | 64 |
| >60 | 6 (20%) | 2 (20%) | 2 (20%) | 1 (17%) | 0 (0%) | 3 (15%) | 14 |
| **Vaccination** |  |  |  |  |  |  |  |
| 1 dose | 6 (20%) | 4 (40%) | 10 (100%) | 0 (0%) | 0 (0%) | 0 (0%) | 20 |
| 2 doses | 14 (47%) | 3 (30%) | 0 (0%) | 1 (17%) | 1 (50%) | 0 (0%) | 19 |
| 3 doses | 10 (33%) | 3 (30%) | 0 (0%) | 5 (83%) | 1 (50%) | 0 (0%) | 19 |

|  | AstraZeneca<br>(n=30) | Pfizer<br>(n=10) | Johnson &<br>Johnson<br>(n=10) | Moderna<br>(n=6) | Mixed<br>(n=2) | Wildtype<br>Infected<br>(n= 20) | Total<br>(n=78) |
| --- | --- | --- | --- | --- | --- | --- | --- |
| **Sex** |  |  |  |  |  |  |  |
| **Male** | 15 (50%) | 2 (25%) | 5 (50%) | 3 (50%) | 0 (0%) | 11 (55%) | 36 |
| **Female** | 15 (50%) | 8 (75%) | 5 (50%) | 3 (50%) | 2 (100%) | 9 (45%) | 42 |
| **Age** |  |  |  |  |  |  |  |
| **>18-60** | 24 (81%) | 8 (75%) | 8 (80%) | 5 (75%) | 2 (100%) | 17 (85%) | 64 |
| **>60** | 6 (19%) | 2 (25%) | 2 (20%) | 1 (25%) | 0 (0%) | 3 (15%) | 14 |
| **Vaccination** |  |  |  |  |  |  |  |
| **1 dose** | 6 (23%) | 4 (38%) | 10 (100%) | 0 (0%) | 0 (0%) | 0 (0%) | 20 |
| **2 doses** | 14 (50%) | 3 (38%) | 0 (0%) | 1 (25%) | 1 (50%) | 0 (0%) | 19 |
| **3 doses** | 10 (27%) | 3 (24%) | 0 (0%) | 5 (75%) | 1 (50%) | 0 (0%) | 19 |

### Pseudovirus production and neutralization assay

The pseudovirus production and assay validation have been described before ^24^. Briefly, a lentiviral expression system was used to produce wildtype and omicron sublineages, EG.5.1, FY.4, BA.2.86, JN.1, and JN.1.4 pseudoviruses. Three plasmids, coding for the MLV-gag/pol backbone, luciferase, and full-length spike protein of the different variants were co-transfected into HEK293T cells using PEI (Polysciences, 24765-1) to produce a single round of infection competent pseudoviruses. The media were changed 24hrs post-transfection and the pseudovirus harvested 72hrs post-transfection. The pseudoviruses were aliquoted and frozen for long-term storage. Virus infectivity of the variants was determined by titration on HEK293T (hACE2-hTMPRSS2)-stable cells and dilution of pseudoviruses giving >20,000RLU was selected for assaying. To test for neutralization, 50 μL of virus was immediately mixed with 50 μL of serially diluted (2×) serum and incubated for 1 h at 37°C. Following the incubation, 10,000 HEK293T (hACE2-hTMPRSS2) cells/well (in 100 μL of media) were directly added to the antibody-virus mixture. The plates were then incubated at 37°C for 72 h. To check for neutralization, HEK293T (hACE2-hTMPRSS2) cells were lysed using a lysis buffer (Promega, E2661). Luciferase intensity was then read on a Luminometer with luciferase substrate according to the manufacturer’s instructions (Promega, E2650). The percentage of neutralization was calculated using the following equation: 100 × (1− (RLU of sample−average RLU of background)/average RLU of virus only − average RLU of background)), where the background was cell-only control. Additionally, as part of the assay controls, a positive control of a pool of convalescent serum from 50 individuals with confirmed COVID-19 and a negative control of a pool of pre-pandemic serum from 50 individuals were included. To determine the ID50 value, a model of the dose-response curve was fit using the sample dilution and the corresponding neuralization percentage.

### Statistical analysis

Data analysis was run on R v4.3.2 and R Studio 2023.12.1. ID_50_ significance was measured using Student’s t-test and Mann-Whitney tests, where significance was considered at p-value < 0.05 (*), p-value <0.001(**), and p-value <0.0001(***). The calculation of ID50 is as described previously ^24^.

## Data Availability

Add data produced are available online at https://doi.org/10.7910/DVN/6DSHMB

## Acknowledgments

We thank all the sample donors for their contribution to the research. We also acknowledge all GISAID data contributors whose genomic sequences and metadata this work is based on.

## Author contribution

Conceptualization and methodology: JN. Investigation: BK, JN, DL, MK, AS, JG, AK, DA, BK, AM, AL, DO, RL, JN. Formal analysis: JN, DL, BK, GG. Resources and funding acquisition: JN, IO, DB, GG, CS, EN. Writing, original draft preparation: DL, JN, BK. Writing, review and editing: all authors

## Ethics declaration

This study was approved by the Scientific and Ethics Review Unit (SERU) of the Kenya Medical Research Institute (SERU ID: 4085 and 4807). Written informed consent was obtained from participants for the collection, storage, and further use of their sample sets in the research.

