## Supplementary Figure 1 & 2 for "Evaluation of population immunity against SARS-CoV-2 variants, EG.5.1, FY.4, BA.2.86, JN.1, and JN.1.4, using samples from two health demographic surveillance systems in Kenya"

**Supplementary Fig. 1 Representation of samples collected for sequencing in Kenyan counties between May 2023 and March 2024.**

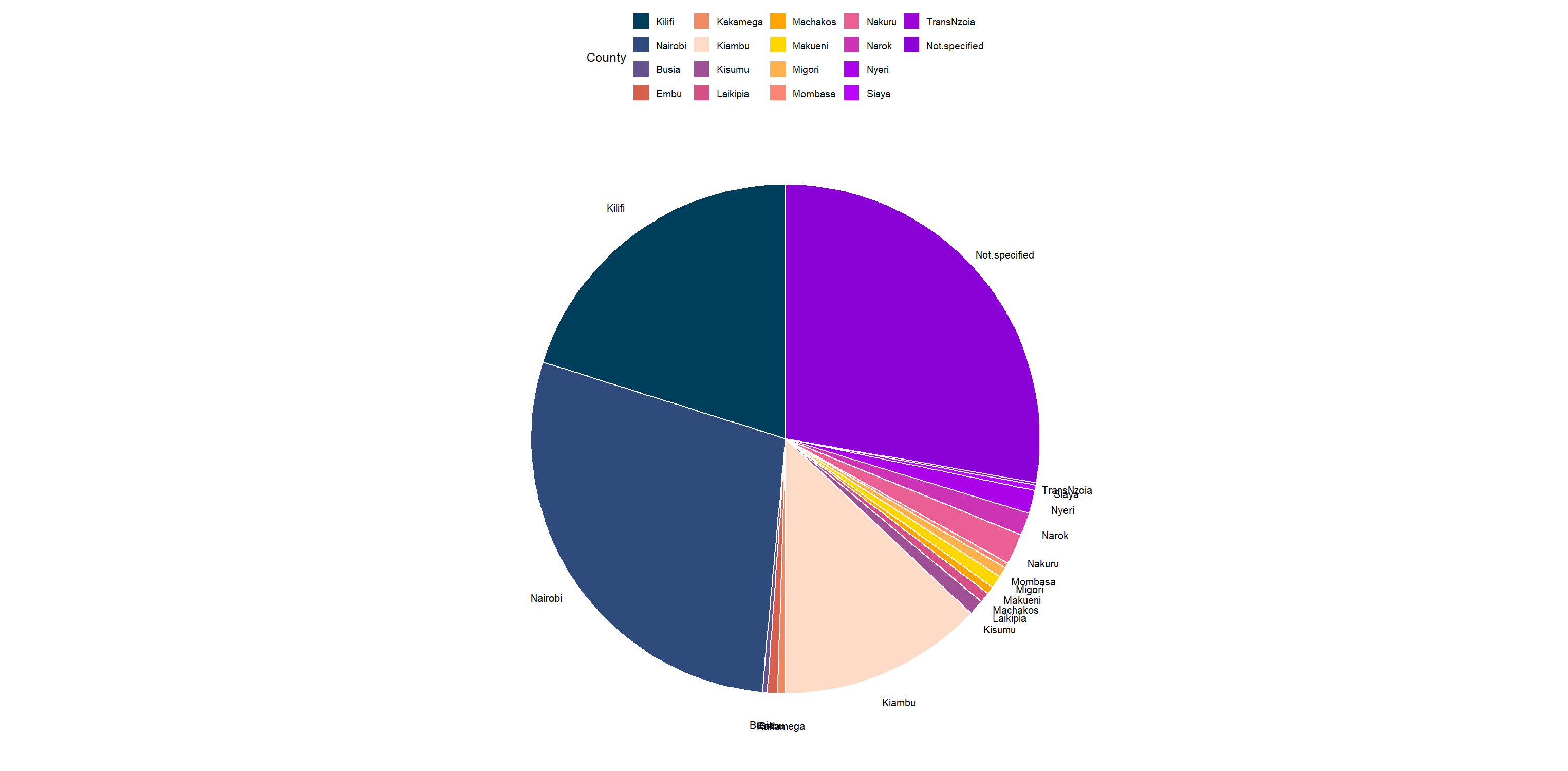

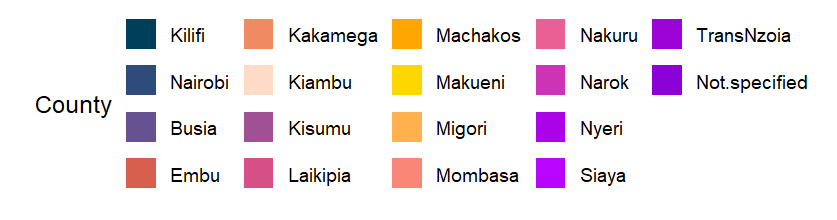

Kilifi

20%

Nairobi

(28%)

Kiambu (13%)

Not Specified

28%

**Supplementary Fig. 1:** The pie chart shows the number of SARS-CoV-2 sequences from samples collected in each county between May 2023 and March 2024. Samples were sequenced in the Kenya Medical Research Institute Wellcome Trust Programme in Kilifi and other centers in Kenya.

**Supplementary Fig. 2: Assessment of neutralization function based on sex and one-dose vaccination in Kenyan population samples***.*

**B.**

**A.**

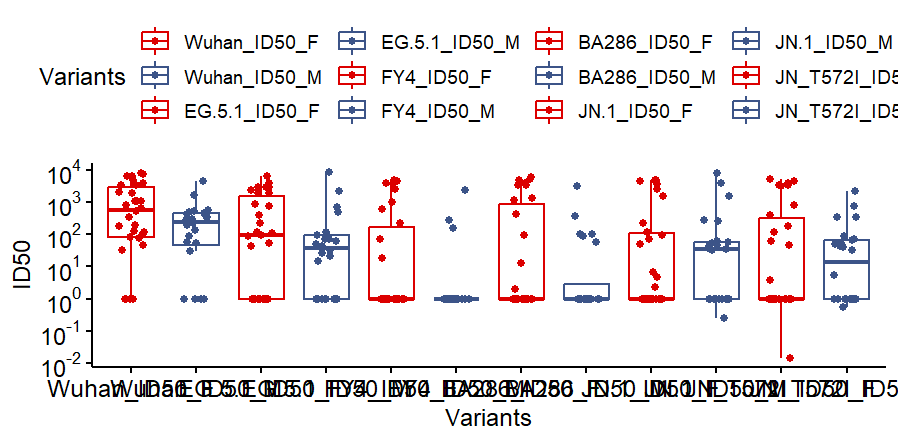

**Female**

**Male**

**Wildtype**

**FY.4**

**BA.2.86**

**logID_50_**

**EG.5.1**

**JN.1**

**JN.1.4**

**ns**

**ns**

**ns**

**ns**

**ns**

**ns**

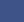

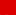

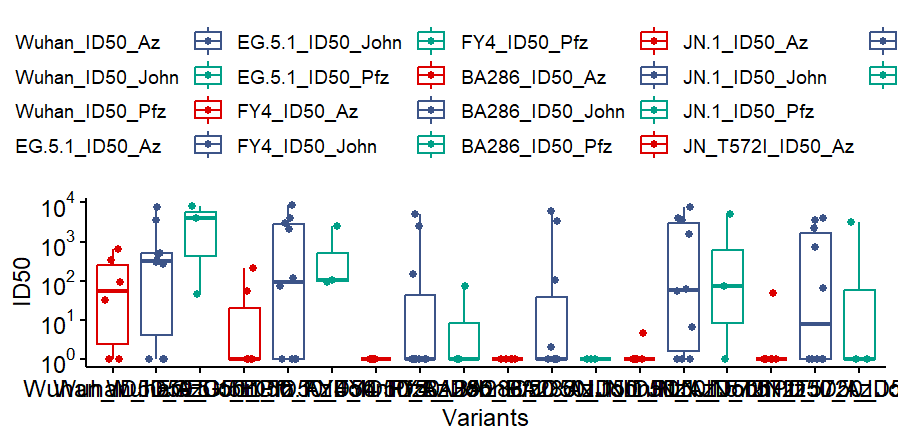

**Wildtype**

**FY.4**

**BA.2.86**

**logID_50_**

**EG.5.1**

**JN.1**

**JN.1.4**

**AstraZeneca**

**Johnson & Johnson**

**Pfizer**

**ns**

**ns**

**ns**

**ns**

**ns**

**ns**

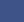

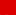

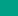

**Supplementary Fig. 2: Assessment of neutralization function based on sex and one-dose vaccination in Kenyan population samples**. A. Shows neutralization function ( log ID_50_ ) against emerging omicron variants in female (n=32) and male (n=23) individuals from our cohort. The legend represents female samples in blue and male samples in red B. Shows neutralization function ( log ID_50_ ) against emerging omicron variants in individuals who received one dose of either Johnson & Johnson (n=10), Pfizer (n=3), or AstraZeneca (n=6) vaccines. The legend colors correspond to samples who received Johnson & Johnson in red, AstraZeneca in blue, and Pfizer in green. Statistical significance was determined by Mann-Whitney tests, and no significance (ns) was detected in all tests.
